# Mixed methods implementation research of oral antiviral treatment for COVID-19 in low and middle-income countries: a study protocol

**DOI:** 10.1101/2024.10.25.24316111

**Authors:** Shanti Narayanasamy, Fiona Gambanga, Caroline E. Boeke, Krishna Udayakumar, Leo Brothers, Cameron R. Wolfe, Chukwuemeka Agwuocha, Maame Nkansaa Asamoa-Amoakohene, Khamsay Dethleuxay, Bridget C. Griffith, Nervine Hamza, Jessica Joseph, Philip Kimani, Robert Kirungi, Norman Lufesi, Nyuma Mbewe, Elizabeth McCarthy, Mwaba Mulenga, Moses Mukiibi, Tamara Mwenifumbo, Lawrence Ofori-Boadu, Ijeoma Okoli, Sompasong Phongphila, Sean Regan, Edson Rwagasore, Alan Staple, Jessica Tebor, Wesley Tomno, Sabine Umuraza, Hayden B. Bosworth, COVID Treatment QuickStart Consortium

## Abstract

**Introduction:** There is an absence of real-world evidence, especially from low- and middle-income countries (LMICs), on the implementation successes and challenges of COVID-19 test and treat (T&T) programs. In 2022, nirmatrelvir/ritonavir was provided as standard of care for mild to moderate COVID-19 treatment in eight LMICs (Ghana, Kenya, Laos, Malawi, Nigeria, Rwanda, Uganda and Zambia). This manuscript describes a research protocol to study novel drug introduction during the COVID-19 health emergency, with implications and learnings for future pandemic preparedness. The goal of the study is to provide simultaneous program learnings and improvements with program rollout, to fill a gap in real-world implementation data on T&T programs of oral antiviral treatment for COVID-19 and inform program implementation and scale-up in other LMICs.

**Methods and analysis:** This multiple methods implementation research study is divided into three components to address key operational research objectives: 1) program learnings, monitoring and evaluation; 2) patient-level program impact; and 3) key stakeholder perspectives. Data collection will occur for a minimum of six months in each country, up to the end of grant. Quantitative data will be analysed using descriptive statistics for each country and then aggregated across the program countries. Stakeholder perspectives will be examined using the Consolidated Framework for Implementation Research implementation science framework and semi-structured interviews.

**Ethics and dissemination:** This study was approved by the Duke University Institutional Review Board (Pro00111388), The study was also approved by the local institutional review boards in each country participating in individual-level data collection (Objectives 2 and 3): Ghana, Malawi, Rwanda and Nigeria. The study’s findings will be published in peer-reviewed journals and disseminated through dialogue events, national and international conferences and through social media.

**Trial registration number:** Clinicaltrials.gov NCT06360783.

**STRENGTHS AND LIMITATIONS OF THIS STUDY:**

1. The knowledge generated though this study will improve understanding of the key characteristics of a strong test and treat (T&T) program, and may translate to improved local T&T program implementation and better patient outcomes.
2. This study uses real-world routinely collected health data and is not designed as a randomised trial; therefore, data quality may be a challenge and the study cannot be used to compare patient outcomes.
3. This study protocol could be adapted to rapidly assess rollout of test and treat interventions to provide real-time learnings in health emergencies.
4. Lessons learned from this study could be applied to other T&T scenarios, particularly around new product introduction, and provide reciprocal innovations to other countries and contexts on the feasibility, challenges, and successes of program implementation.
5. This study is designed to fill a gap in real-world implementation data on T&T programs of oral antiviral treatment for COVID-19 and inform program implementation and scale-up in low- and middle-income countries.

## INTRODUCTION

The COVID-19 pandemic has been a harsh reminder of the extensive harm a new pathogen can cause globally. As of March 2024, four years since its emergence, SARS-CoV-2 has caused an estimated 774 million cases and 7 million deaths globally,^1^ exposing significant global inequities in access to vaccines, therapeutics and other critical response interventions.^2-5^ The estimated likelihood of experiencing a pandemic similar to COVID-19 in one’s lifetime is currently approximately 38%, and is predicted to double in the coming decades.^6^ The COVID-19 pandemic has demonstrated that therapeutics are a key partner to vaccines and diagnostics in limiting mortality among vulnerable individuals.

In December 2021 the US Food and Drug Administration granted Emergency Use Authorization status to two oral antiviral treatments for COVID-19: nirmatrelvir boosted with ritonavir (Paxlovid) developed by Pfizer, and molnupiravir (Lagevrio) developed by Merck. Nirmatrelvir/ritonavir reduces mortality and decreases hospitalisations in unvaccinated, non-hospitalised adult patients at high risk of progression to severe COVID-19 disease.^7^

Both nirmatrelvir/ritonavir and molnupiravir have entered the market in most high-income countries (HICs). The most recent data up to May 2023 shows, however, the majority of the over 68 million courses purchased were procured by HICs (70%) and global entities (27%), with just 1% purchased by upper-middle income countries and 2% by lower-middle income countries.^8^ Low income countries have relied on global entities such as UNICEF, Global Fund and the European Union Preparedness and Response Authority for antiviral supply.

There is an absence of real-world evidence, especially from low- and middle-income countries (LMICs), on how oral antivirals perform outside of stringent clinical trials settings – where patients may not receive treatment within the recommended five days of symptom onset, or patients have greater immunity from vaccination or natural infection.

Despite these challenges, LMICs have been actively working toward locally adapted COVID-19 test and treat (T&T) solutions to protect those most vulnerable to severe disease, older individuals, and those with underlying health conditions. Local solutions using oral antivirals may lower the burden of severe and critical COVID-19 management on health personnel and services, while building capacity within primary care to respond to future emerging infections.^9-11^ T&T approaches are familiar to many LMIC contexts due to HIV and malaria programs, and are undergird by two key foundations: widely available, rapid and decentralised testing, and affordable, accessible treatment that can be deployed at a primary healthcare level.^12 13^

From late 2022 to 2023, nirmatrelvir/ritonavir was introduced as standard of care in eight LMICs (Ghana, Kenya, Laos, Malawi, Nigeria, Rwanda, Uganda and Zambia) in partnership with the COVID Treatment QuickStart Consortium. QuickStart brings together Duke University, Americares, the Clinton Health Access Initiative, and the COVID Collaborative as implementing partners, with support from the Open Society Foundations, Pfizer, and the Conrad N. Hilton Foundation, to work in partnership with LMICs in Africa and Asia. In partnership with QuickStart, countries obtained local regulatory approval for nirmatrelvir/ritonavir, procured treatment for deployment to treatment sites, and supported training of health facility staff in T&T approaches. Treatment sites were public health facilities providing COVID-19 testing services (at the discretion of each country), and private facilities and test locations that provide services free to participants. These included community-level health services (district hospital, health centres, health posts), provincial hospitals and central hospitals.

This manuscript describes a research protocol to study novel drug introduction during the COVID-19 health emergency, with implications and learning for future pandemic preparedness. In this study, we will aim to describe the implementation processes, patient outcomes, stakeholder perspectives, successes, and challenges of the COVID-19 oral antiviral T&T program in the eight LMICs partnered with QuickStart. The goal of the study is to provide simultaneous program learnings and improvements with program rollout, to fill a gap in real-world implementation data on T&T programs of oral antiviral treatment for COVID-19 and inform program implementation and scale-up in other LMICs.

## METHODS AND ANALYSIS

### Conceptual framework and key research questions

Almost eighty percent of the world’s population live in LMICs,^14^ and age-stratified infection fatality rates from COVID-19 are estimated to be approximately two times higher in LMICs compared with HICs.^15^ Country and context specific factors such as gross domestic product, population density, urbanisation, seasonality, interpersonal and government trust, and access to vaccination have driven different rates of infection and mortality.^16-18^ Country-based and context-based implementation research is essential to develop innovative and tailored approaches for a successful COVID-19 T&T program, and to ultimately ensure successful scale up.

The study seeks to answer the following questions:

1. What are the operational considerations for introducing COVID-19 T&T programs in LMICs?
2. What high-risk populations and entry points are feasible to target for T&T implementation in LMICs?
3. What impact do COVID-19 T&T programs have on the number of high-risk patients identified, time to treatment initiation and the total cost per case initiated?
4. Are T&T programs considered feasible and acceptable by key stakeholders including Ministry of Health (MoH), public sector personnel, health care workers and patients?

### Objectives

The COVID-19 T&T program has been implemented in eight LMICs (Ghana, Kenya, Laos, Malawi, Nigeria, Rwanda, Uganda and Zambia) who will contribute monitoring and evaluation data, and programmatic data for a minimum of six months (Objective 1). Additionally, a multiple methods implementation research study, conducted in partnership with the Ministry of Health in five countries (Ghana, Malawi, Nigeria, Rwanda, and Zambia) will be conducted (**Figure 1**). The implementation research is divided into two components to address the key operational research objectives: Patient-Level Program Impact (Objective 2) and Key Stakeholder Perspectives (Objective 3). Each country (Ghana, Malawi, Nigeria, Rwanda and Zambia) will collect data for a minimum six month period, up to the end of the grant. The five countries conducting implementation research opted in to this part of the study based on the preference of local Ministries of Health. The implementation timeline of each component is detailed in **Figure 2**.

**Figure 1:**
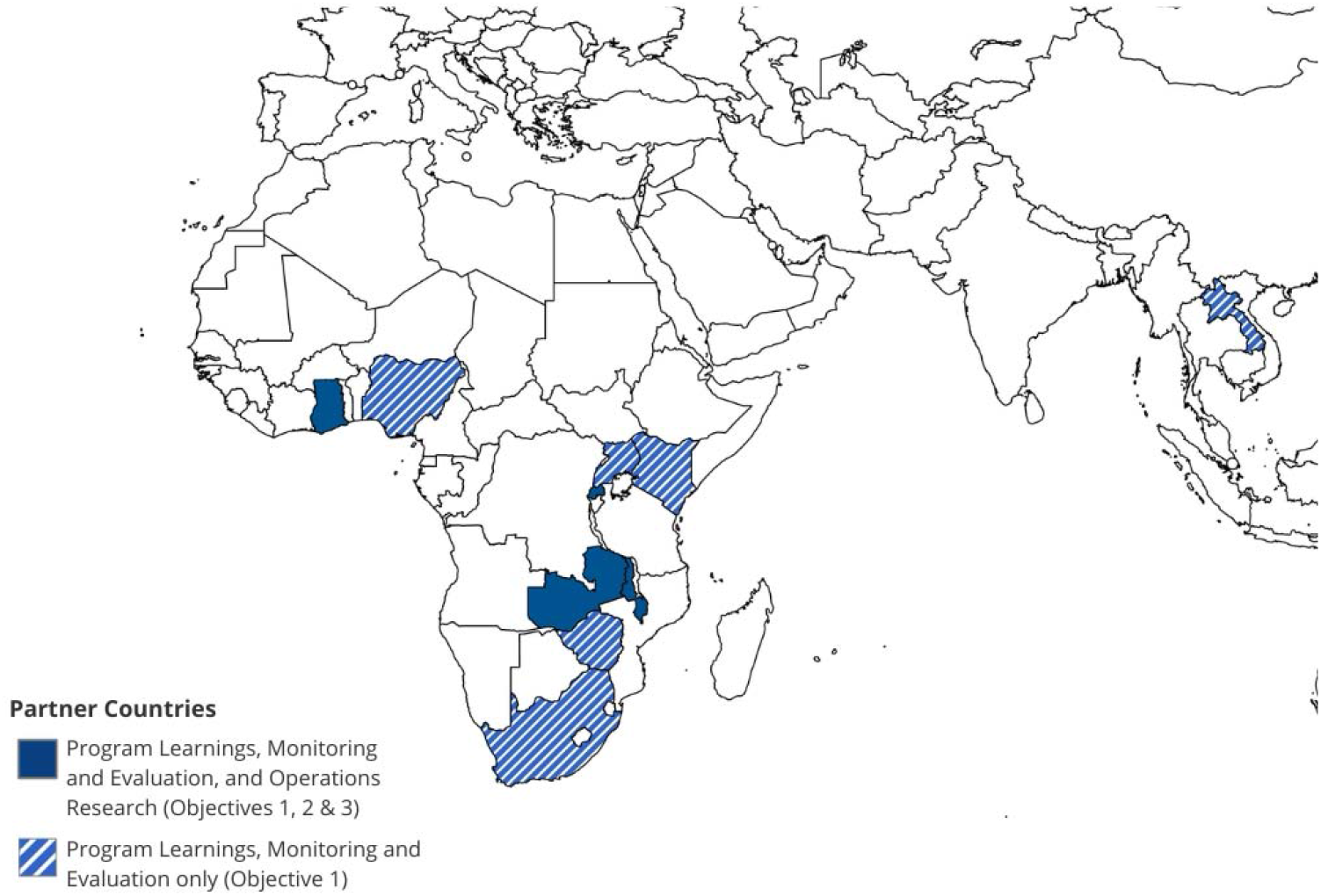
Map of participating countries

**Figure 2:**
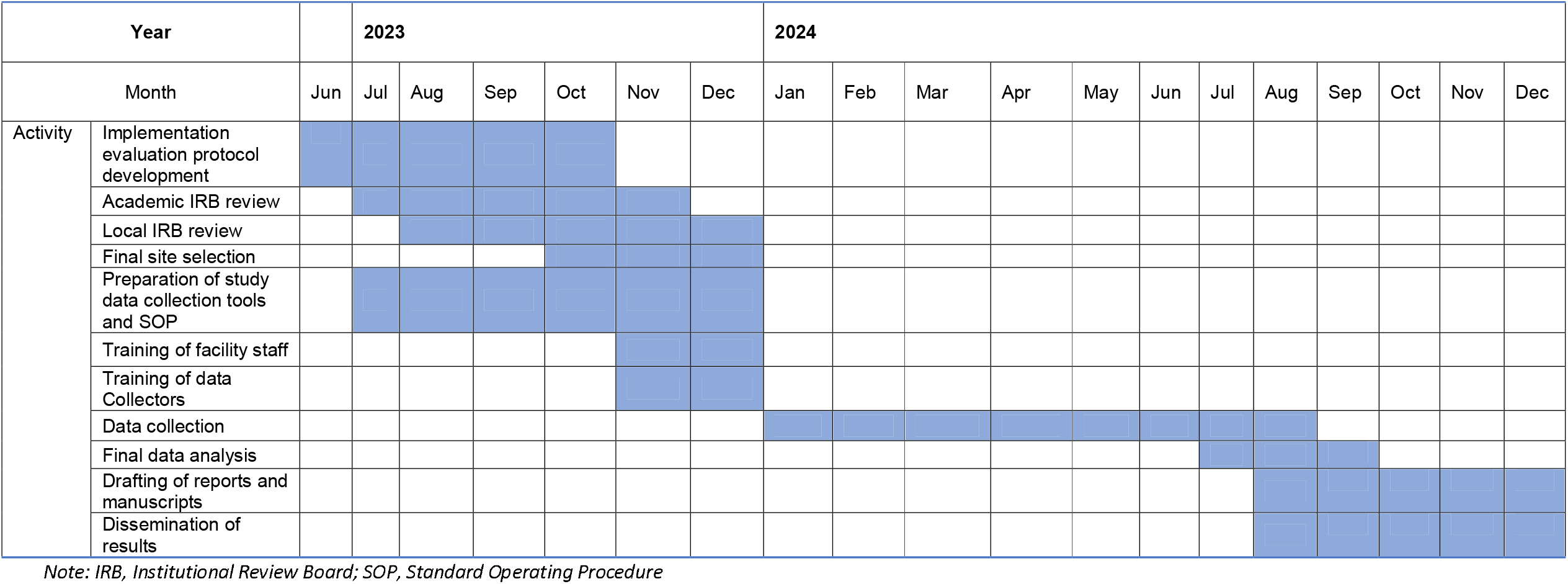
Timeline of study implementation

1. **Program Learnings, Monitoring and Evaluation**: To identify contextual factors impacting the implementation and deployment of the COVID-19 T&T model and the cost per case initiated under the T&T model (all eight countries, Ghana, Kenya, Laos, Malawi, Nigeria, Rwanda, Uganda and Zambia).
2. **Patient-Level Program Impact**: To measure key service delivery metrics among those testing positive for SARS-CoV-2 at high-risk of severe COVID-19 illness (Ghana, Malawi, Nigeria, Rwanda and Zambia).
3. **Key Stakeholder Perspectives**: To understand the perspectives of key stakeholders, including MoH and public sector personnel, health care workers and patients on the feasibility and acceptability of the COVID-19 T&T program (Ghana, Malawi, Nigeria, Rwanda and Zambia).

### Objective 1: Program Learnings, Monitoring and Evaluation

This objective aims to identify contextual factors impacting the implementation and deployment of the COVID-19 T&T model in eight LMICs (Ghana, Kenya, Laos, Malawi, Nigeria, Rwanda, Uganda and Zambia). This objective includes documentation of national COVID-19 T&T policies and guidelines, and identification of key aspects of adoption and implementation of the T&T program. This objective will also describe country-level metrics around distribution of COVID-19 treatment, provider and administrator organisational readiness to implement the program and cost per case initiated under the current model.

### Study Design

Study monitoring of the implementation of the T&T program will leverage existing national MoH routine monitoring and evaluation (M&E) program data at the aggregate level. Data collection will be retrospective on a monthly basis for a minimum of the first six months of the program. Additionally, activities conducted to prepare for program implementation and to identify implementation gaps will be documented.

### Inclusion Criteria

All treatment sites followed established country protocols for COVID-19 patient testing and treatment with oral antivirals based on international guidelines (**Supplemental 1, Figure 1**).^19^ Patients testing positive for SARS-CoV-2 were offered oral antiviral treatment if: >12 years old, mild/moderate COVID-19 disease, within 5 days of symptom onset, at high-risk for progressing to severe COVID-19 infection, and without contraindications for oral antiviral treatment. The high-risk population included: age ≥50 years, hypertension, diabetes, cardiac disease, chronic lung disease, chronic liver disease, cerebrovascular disease, dementia, mental health disorders, chronic kidney disease, disability, immunosuppression, obesity (BMI >30kg/m2), tuberculosis, pregnancy, smoking and cancer. Small variations in high-risk conditions were permitted between countries due to local risk factors. High-risk conditions were determined through self-report and review of patient medical records. Antiviral treatment was provided as standard of care and free to the patient at all treatment sites across the eight countries.

### Site Selection & Sample Size

The number of treatment sites varies by country (**Table 1**). Treatment sites were determined by MoHs as existing sites with the capacity to conduct COVID-19 testing and prescribe oral treatment. All treatment sites in the eight T&T countries will be included in Objective 1 data collection.

**Table 1:**
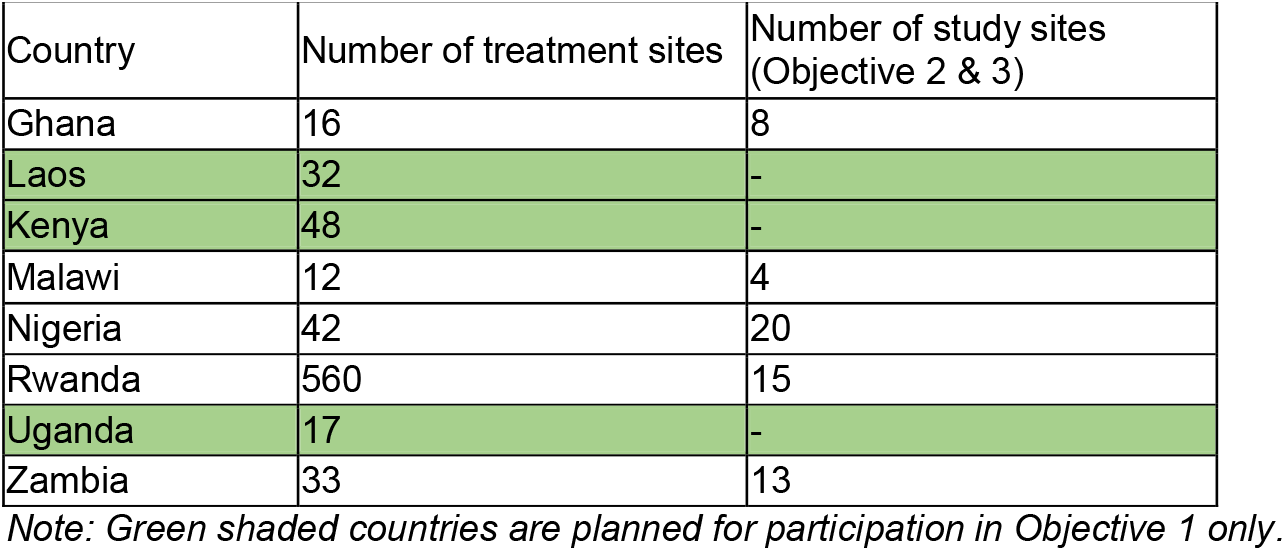
Estimated number of activated sites and study sites for the COVID-19 T&T program per country.

### Data collection

#### Programmatic monitoring

Countries will initiate data collection according to when they activate their T&T programs, recognising start times will vary within, and between, countries.

Countries will submit five types of data:

1. *Aggregate service delivery data*. Collected at the national level, and where possible, at the facility level. Data will include the number of COVID-19 tests conducted, the number of positive COVID-19 tests and the number of nirmatrelvir/ritonavir prescriptions (**Supplemental 2, Facility Monthly Summary**).
2. *Facility information data*. Collected at the beginning of the program through a questionnaire. Includes descriptive information and key characteristics of each participating facility (**Supplemental 1, Figure 2**). Data may also be collected from site visits or supportive supervision reports to monitor the progress of program implementation.
3. *Implementation progress data*. Collected on the preparation and implementation activities conducted for the COVID-19 T&T program on topics such as policy, supply chain, training, quality assurance and data systems will be reported by each country at the beginning of the program and towards the end of implementation. Data may also be available from site visits or supportive supervision reports to monitor implementation progress.
4. *Cost data*. Data on program costs including commodities, training, staff time, printing of registers and logbooks.
5. *Adverse event data*. Data will be collected on severe adverse events (SAE). Any SAE from a participant receiving oral antiviral treatments for COVID will be reported to the manufacturer within 24 hours of the study team becoming aware of the event. Data will also be collected on the presence and continuous functioning of a local recording and reporting system for SAEs.

#### Organisational Readiness for Implementing Change survey administration

At baseline, we will administer the 12-item Organizational Readiness for Implementing Change (ORIC) measure, based on the ability of staff to initiate a program, put forth greater effort, and be persistent and cooperate with one another to implement the program.^20^ Before the ORIC instrument is administered, we will collect a Pre-Implementation Survey including information about the organisation, the role of the persons at the organisation completing the survey and that person’s characteristics (**Supplemental 1, Figure 3**). This survey will help characterise organisational readiness and identify health organisation change efforts that do not achieve desired benefits.

### Data analysis plan

All quantitative data will be analysed using descriptive statistics for each country and then aggregated across the program countries. Site information will be summarised, and key descriptors and themes drawn out in summary reports. Implementation data will be analysed by examining key themes and takeaway learnings, and then summarising them by country and overall program. Data analysis will take place using Excel, SAS, and Stata software.

### Objective 2: Patient-Level Program Impact

The aim of this objective is to understand the impact of the program at the patient level in five countries (Ghana, Malawi, Nigeria, Rwanda and Zambia). Country MoHs opted-in to participation in Objectives 2 and 3. We will measure the following key metrics among those testing positive for SARS-CoV-2: the number of patients at high-risk for severe COVID-19 disease, the number eligible for oral antiviral treatment, the number testing positive for SARS-CoV-2 within 5 days of onset, the uptake of oral antiviral treatment and treatment completion among eligible patients, treatment side effects, and the reasons for not commencing treatment among those eligible, or reasons for not completing treatment among those prescribed.

### Study Design

Facility registers or facility paper-based forms will be introduced as part of the COVID-19 T&T program at study sites (**Supplemental 2, Patient Register)**. Data points will be collected uniformly from all Objective 2 countries through these standardised registers/forms. Patient-level data will be retrospectively collected for a minimum six month period, beginning at the commencement of the T&T program in each country, up to June 2024.

### Site Selection

From the treatment sites in each country, we will use purposeful sampling to generate a list of ‘study sites’ to implement Objectives 2 and 3. Sites will be selected in collaboration with MoHs in each country based on diversity within the sample, for example, rural and metropolitan sites, geographic representation, caseload, and representation of different patient populations. At a minimum, four study sites will be selected per country (**Table 1**). This sample cannot be considered nationally representative in a statistical or proportional sense, but rather aims to represent a wide range of experiences of implementing the COVID-19 T&T program. In assessing the diversity of the sample, the following characteristics will be examined: the population being targeted, caseload, epidemic context, rural and metropolitan location, and geographical region.

### Patient selection

All patients who test positive for SARS-CoV-2 at the study site during the data collection period of the study will be included. Data collection will occur retrospectively from the health facility and will not involve direct interaction with patients. As such, informed consent will not be sought. Rather, we will not collect any personal identifying information gathered from the facility data source during the data extraction.

### Sample Size

The sample size of COVID-19 positive, high-risk individuals targeted for this study is at least 200 per country. The sample size was determined to ensure precision when estimating the proportions for multiple binary endpoints. For example, the proportion of treated patients who have known high-risk factors. The sample size estimation depends on the level of precision desired, as reflected in the width of confidence intervals. For estimation of a binary proportion, a confidence interval width of 0.10 would correspond to a confidence interval from 0.85-0.95, for example. We determined that a sample size of 200 would preserve a confidence interval width <0.15 for any true proportion ranging between 0.6 and 0.9.

### Data Collection

Individual-level data will be collected retrospectively using facility registers or facility paper-based forms as described above. These data include presenting symptoms, eligibility for oral antiviral treatment, and provision of treatment. Health care workers are requested to follow-up with patients at 10 and 30 days post treatment initiation either in person in clinic, or with a phone call; these follow-up data will be collected. In addition, data from routine laboratory information management systems, DHIS-2, or electronic medical record systems in use at sites or the national level will be reviewed if they capture study indicators of interest.

All data collectors will receive training that includes the objectives and design of the study, use of the data collection tools, and research ethics. Data will be collected electronically using SurveyCTO^®^ by data collectors and uploaded via encrypted forms to a secure, encrypted server. The data access will be governed by local guidelines.

### Data analysis plan

Patients’ demographic characteristics and baseline clinical risk factors, severity of disease and other relevant characteristics will be reported using descriptive statistics. Continuous data will be summarised by measures including the mean, standard deviation, median, first and third quartiles, minimum, and maximum. Categorical data will be presented by frequencies (n and percentage) or contingency tables. Primary and secondary outcomes for cases initiating treatment will be reported using similar descriptive statistics. If applicable, multivariable analyses will be conducted using regression models (e.g. logistic regression, general linear model, negative binomial) based on the distribution of the measure. Practice variation and patient clustering may be examined using modelling and missing data will be handled with techniques such as single imputation or Markov chain Monte Carlo data augmentation algorithm.

Unless otherwise noted, 2-sided p-values <0.05 will be considered statistically significant. Statistical analyses of the aggregate, de-identified data will be performed using SAS software (version 9.4 or later; SAS Institute, Cary, NC) and R (R Core Team, 2021).

### Objective 3: Key Stakeholder Perspectives

The aim of this objective is to understand the perspectives of key stakeholders on the COVID-19 Test and Treat program. Objective 3 will be assessed through qualitative interviews of key stakeholders in five countries (Ghana, Malawi, Nigeria, Rwanda and Zambia). We will conduct in-depth, semi-structured interviews with individuals from the following stakeholder groups: 1) MoH or public sector personnel, 2) healthcare workers involved in the COVID-19 T&T programs; 3) patients who received care from the COVID-19 T&T program.

### Study Design

The Consolidated Framework for Implementation Research (CFIR) is a meta-theoretical implementation science framework used to describe heterogeneity in implementation across settings, as well as the relative effect of key determinants in influencing implementation outcomes.^21^ The CFIR has been widely used to examine barriers and facilitators to program implementation and is the structural framework used to guide the qualitative aspects of the this study.

### Sample Selection and Sample Size

Healthcare workers and patients will be drawn from study sites through convenience sampling. Healthcare workers will be included if involved in administering COVID-19 testing or treatment at their site. Patients will be included if they are ≥18 years old, test positive for SARS-CoV-2 and were offered oral antiviral treatment (regardless of whether treatment was initiated or completed). MoH/public sector personnel will be purposively sampled within each country to identify and select participants with a diversity of perspectives and experiences, and familiarity with the COVID-19 T&T program.^22^ MoH/public sector personnel will be considered for participation if employed within the MoH national, provincial, district or public sector and involved in developing or implementing the COVID-19 T&T program. A record will be maintained of how many key stakeholders decline to participate and their reason for declining in order to better understand potential biases. Participants are free to decline participation without providing a reason.

The sample size will be driven by thematic saturation and qualitative power.^23^ Based on previous work, our target sample size for the number of completed interviews is approximately 10-12 per stakeholder group per country (n=30-36 per country).^24^ Prior research suggests that a median of 8 to 16 in-depth interviews is needed to reach 80% and 90% saturation, respectively.^25^

### Data Collection

MoH/public sector personnel interview guides will address policy considerations for implementation of the program (**Supplemental 1, Figure 4**). Healthcare worker interview guides will address facility-based barriers and challenges in implementation of the program (**Supplemental 1, Figure 5**). Patient interview guides will explore the patient experience receiving care from the COVID-19 T&T program (**Supplemental 1, Figure 6**).

Written or verbal informed consent will be sought prior to conducting interviews. Interviews may be conducted in person or remotely using programs such as Zoom or WhatsApp. Interviews will begin within three months of the official launch of the program in each country, depending on the local implementation timelines.

Qualitative data collectors will be trained by experienced qualitative researchers either directly or in a ‘train-the-trainer’ model. In this model qualitative researchers will train local data collectors and/or trainers on qualitative research methodology, interviewing skills, note-taking skills, audio recording and obtaining informed consent. These trainers in turn will train local data collectors prior to data collection, with the support and feedback of the qualitative researchers. Trained data collectors will administer the interview guides and audio record the interviews using SurveyCTO^®^. Following each interview, note-taking forms will be completed by the data collectors, recording and summarising the interview. Note-taking forms and audio interviews will be uploaded to a secure, encrypted server for analysis.

### Data Analysis Plan

We will analyse the qualitative interview data following a rapid content analysis technique,^26-29^ using Microsoft Excel version 2019 to support coding and analysis. At least two researchers will review all note-taking forms to develop interview summary templates or debrief notes using content analysis guided by CFIR. Rigor and validity will be established by independently coding and summarising the data, discussing emerging codes and thematic groupings during meetings, and receiving input from a third researcher to resolve any discrepancies. Following a systematic assessment of context, barriers and facilitators identified from the qualitative interviews, we will use the CFIR-Expert Recommendations for Implementing Change (ERIC) implementation strategy match tool, which will provide us with a list of implementation strategies (e.g., facilitation, audit, and feedback, develop quality monitoring systems, etc.) to consider for each country. These strategies will shared with program administrators and local MoHs in each country. This feedback loop will allow for real-time programmatic change to better suit local populations and needs.

## DISCUSSION

The system improvement learnings generated by this research has the potential to strengthen and solidify the standard of care for mild to moderate COVID-19 through T&T programs in the eight LMICs where the program is being implemented (Ghana, Kenya, Laos, Malawi, Nigeria, Rwanda, Uganda and Zambia). The knowledge generated though this study will improve understanding of the key characteristics of a strong T&T program and may translate to improved local T&T program implementation and better patient outcomes. These direct patient-level impacts include decreased hospitalisation and mortality from COVID-19, and improved uptake of COVID-19 testing through demand generated by treatment availability.

The evaluation approach of the three study objectives work synergistically such that the overall impact of the study may support partner MoH countries to be able to apply lessons learned during implementation of this program to other T&T programs, supporting evaluation of future products/programs for emerging diseases. Through program learnings and feedback from stakeholders, the study also has the potential to support integration of the COVID-19 T&T program into existing health programs such as HIV and TB as a path to sustainability, and encourage pandemic preparedness through syndromic diagnosis and management that may be rapidly pivoted to future novel pathogens.

A challenge of this study design, which includes retrospective analysis of observational patient data, is that program implementation will change over time, as will the epidemiologic context of the COVID-19 epidemic. Thus, it will be important to carefully document and analyse program changes over time. We will ensure that study teams document relevant contextual and policy changes and any other factors that impede or promote SARS-CoV-2 testing, e.g., reagent stock outs, refresher training, and the availability of treatments and vaccines.

Another limitation of the study is the variable quality of data in activated and study sites, especially for a new disease program. The variability in quality of data will be somewhat mitigated through site supervision and support, as well as providing support for improvements in data management solutions. We will make attempts to standardise data collection procedures in facility registers or paper-based forms where possible.

This study protocol provides investigators with a template that can be rapidly adapted to assess the implementation of other test and treat interventions, with the potential to provide real-time learnings in future health emergencies.

### Patient and Public Involvement

No patients or members of the public were involved in the design or development of this protocol. The study was developed during a phase of the COVID-19 pandemic where morbidity and mortality from COVID-19 was high and urgent provision of oral treatment to LMICs was a priority. As a result, full community and patient consultation was not undertaken. However, the study was designed to provide rapid learnings and feedback at a country level to better adapt the program to local needs.

## Data Availability

All data produced in the present study are available upon reasonable request to the authors

## ETHICS AND DISSEMINATION

This study was approved by the Duke University Institutional Review Board (Pro00111388), The study was also approved by the local institutional review boards in each country participating in individual-level data collection (Objectives 2 and 3): the Ghana Health Services Ethics Review Committee (017/11/22), the National Health Sciences Research Committee, Malawi (#23/03/4025), the Rwanda National Research Ethics Committee (105/RNEC/2023), the Institutional Review Board ERES Converge (Zambia) (23-Jan-023) and the National Health Research Ethics Committee in Nigeria (NHREC/01/01/2007-11/01/2024). The study’s findings will be published in peer-reviewed journals and disseminated through dialogue events, national and international conferences and through social media.

## AUTHORS’ CONTRIBUTIONS

SN and FG contributed equally to this paper. *Conception and design of the study*: KU, CW, HB, CB, KU, LB, SN, EM, BG, JJ, JT, NH and FW. *First draft of the manuscript*: SN, FG. *Critical revision of the manuscript for important intellectual content:* All authors. All authors take responsibility for the accuracy of the discussion and had authority over manuscript preparation and the decision to submit the manuscript for publication.

COVID Treatment QuickStart Consortium members: *to be listed depending on journal requirements*.

## PATIENT CONSENT FOR PUBLICATION

Not applicable.

## SUPPLEMENTAL MATERIAL

(see separate file)

