## Supplemental 1 for "Mixed methods implementation research of oral antiviral treatment for COVID-19 in low and middle-income countries: a study protocol"

**SUPPLEMENTAL MATERIAL**

**CONTENTS**

**Figure 1:** Covid-19 Oral Antiviral Outpatient Initiation Algorithm	Page 2

**Figure 2:** Facility Information Sheet	Page 4

**Figure 3:** Pre-Implementation Survey	Page 8

**Figure 4:** MoH/Public Sector Personnel Interview Guide	Page 9

**Figure 5:** Healthcare Worker Interview Guide	Page 10

**Figure 6:** Patient Interview Guide	Page 11

**Figure 1: Covid-19 Oral Antiviral Outpatient Initiation Algorithm**

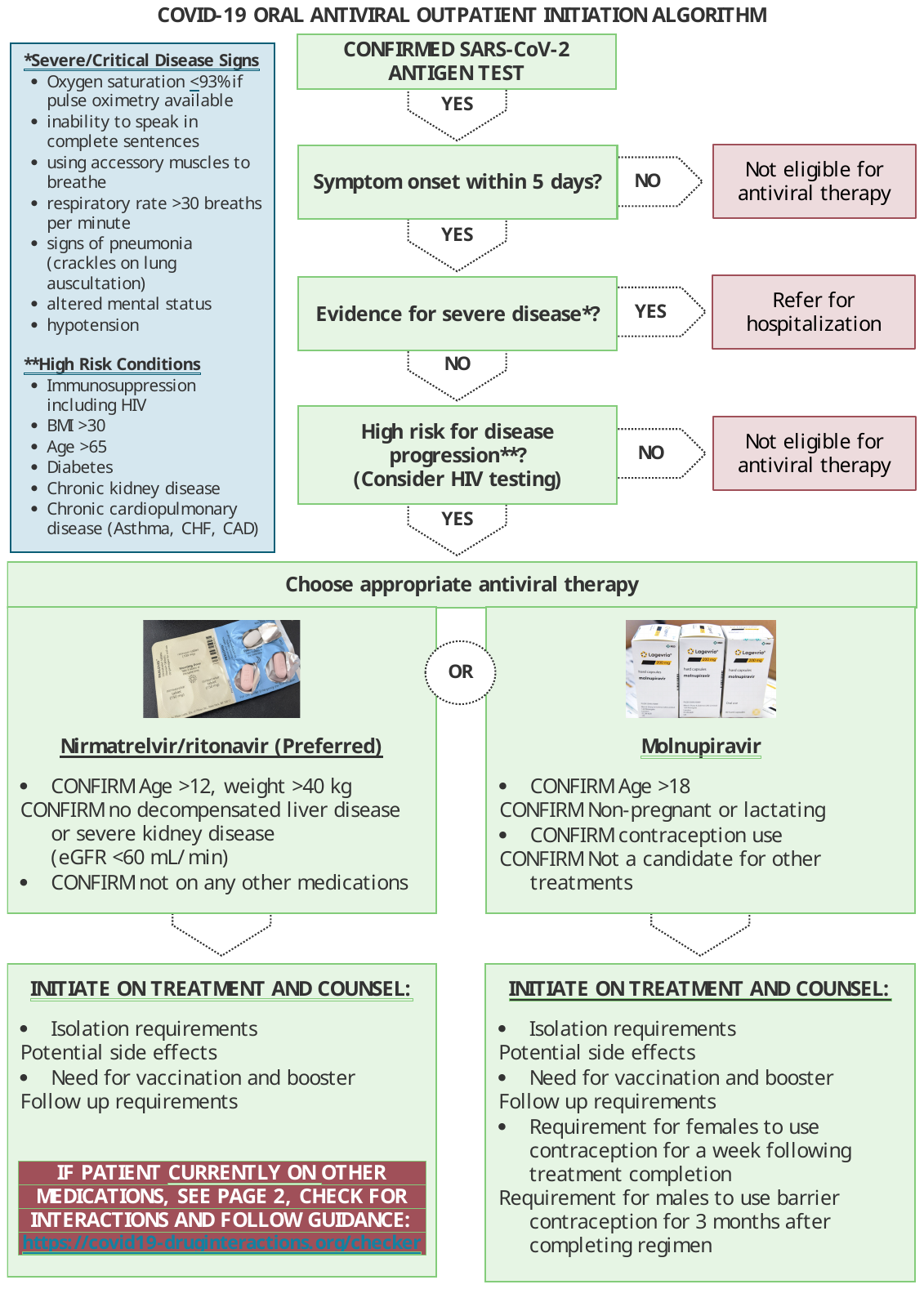

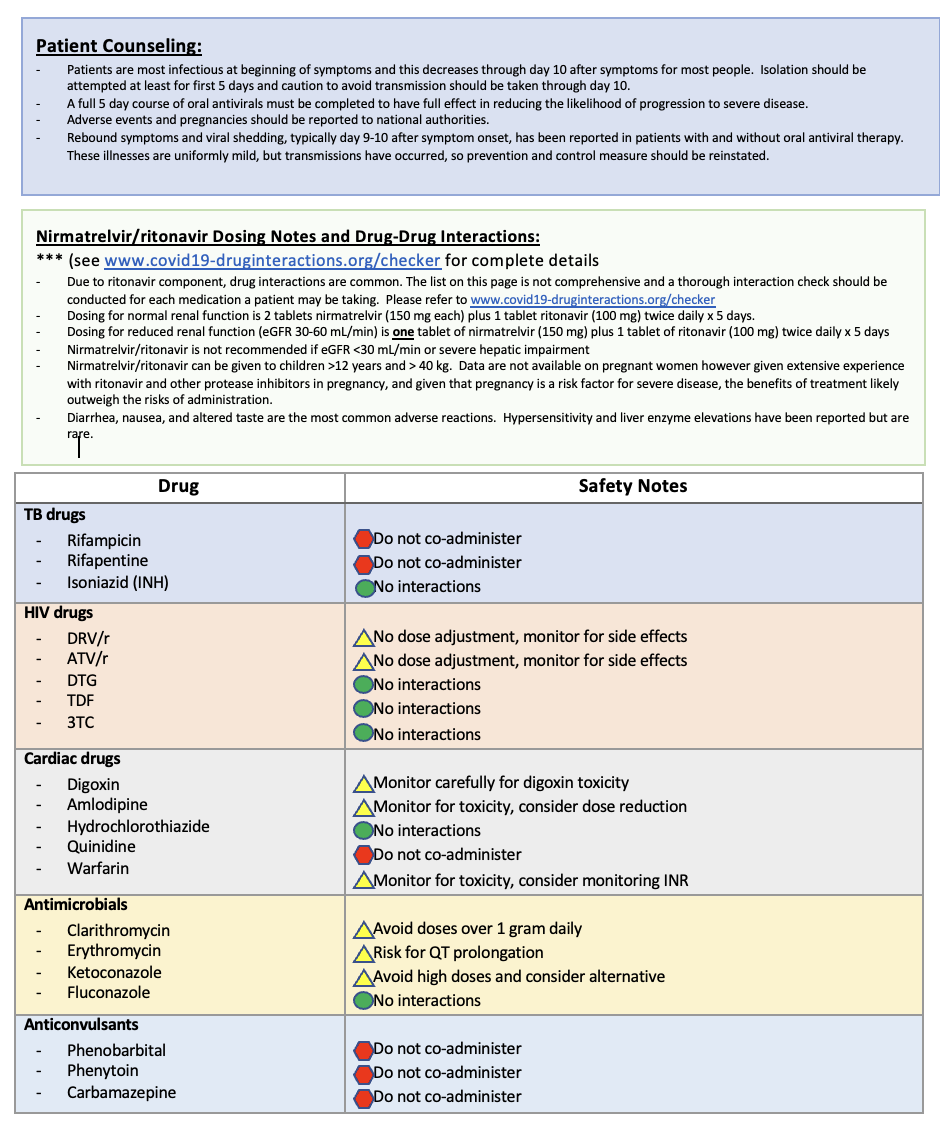

**Figure 2: Facility Information Sheet**
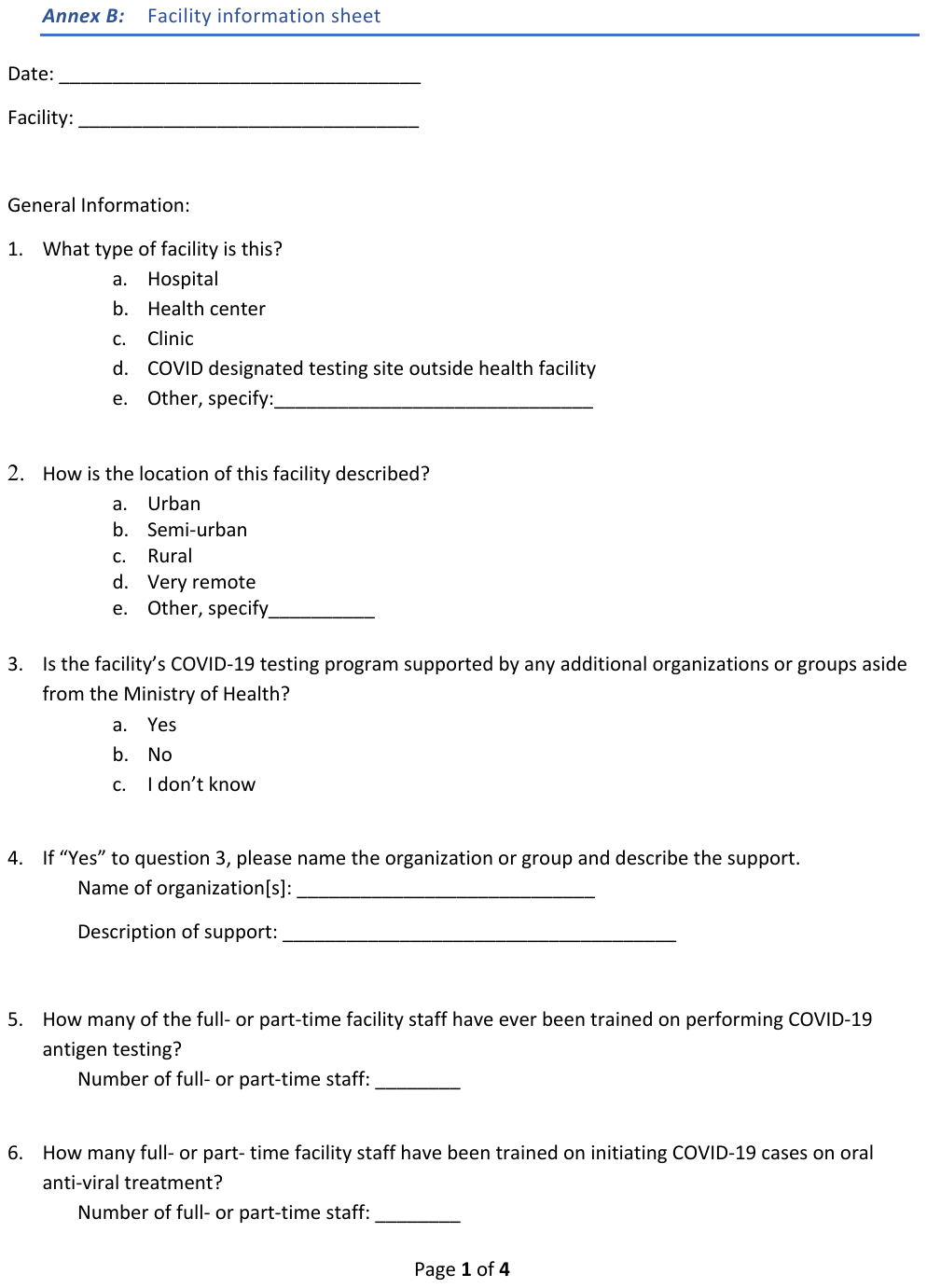

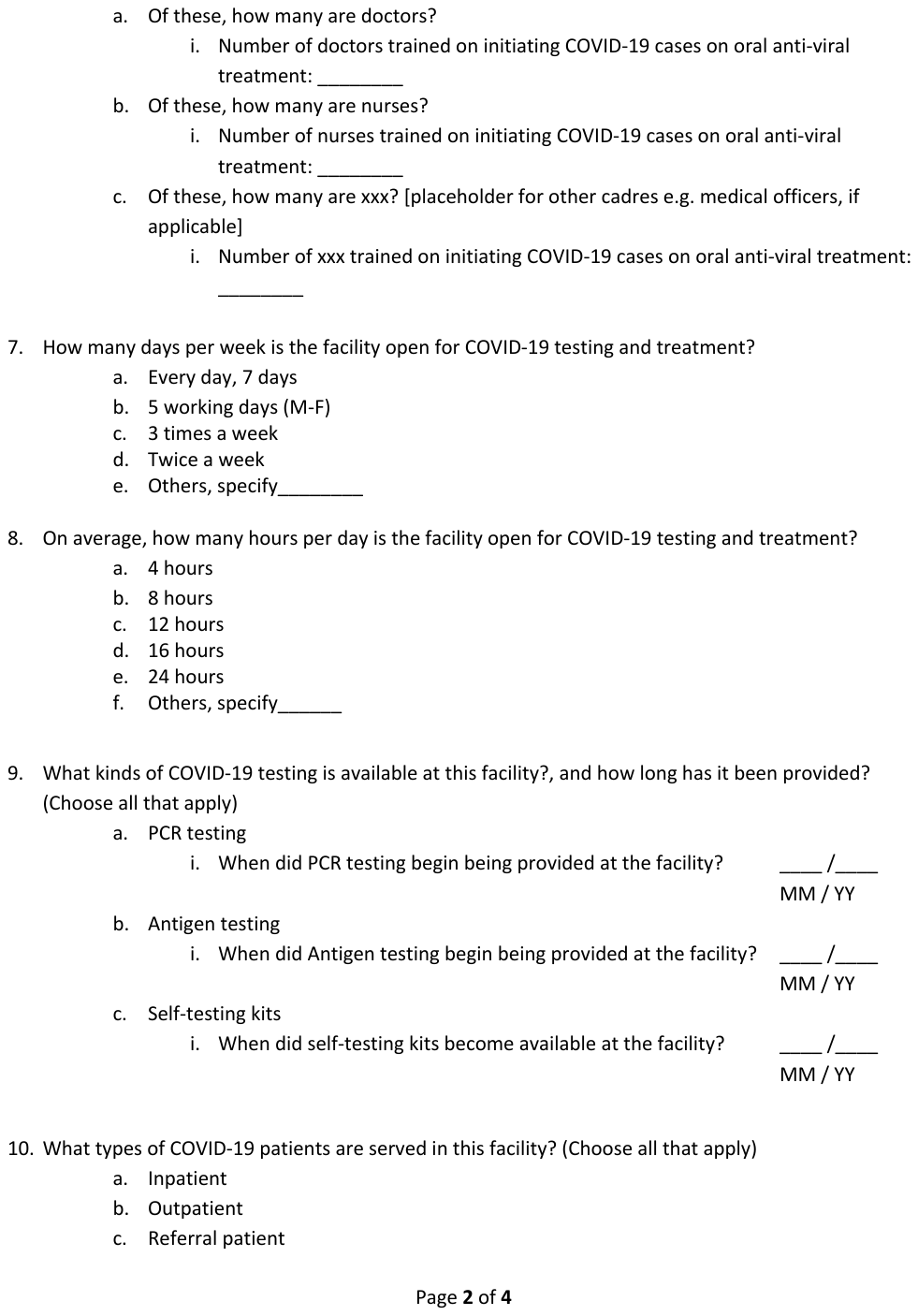

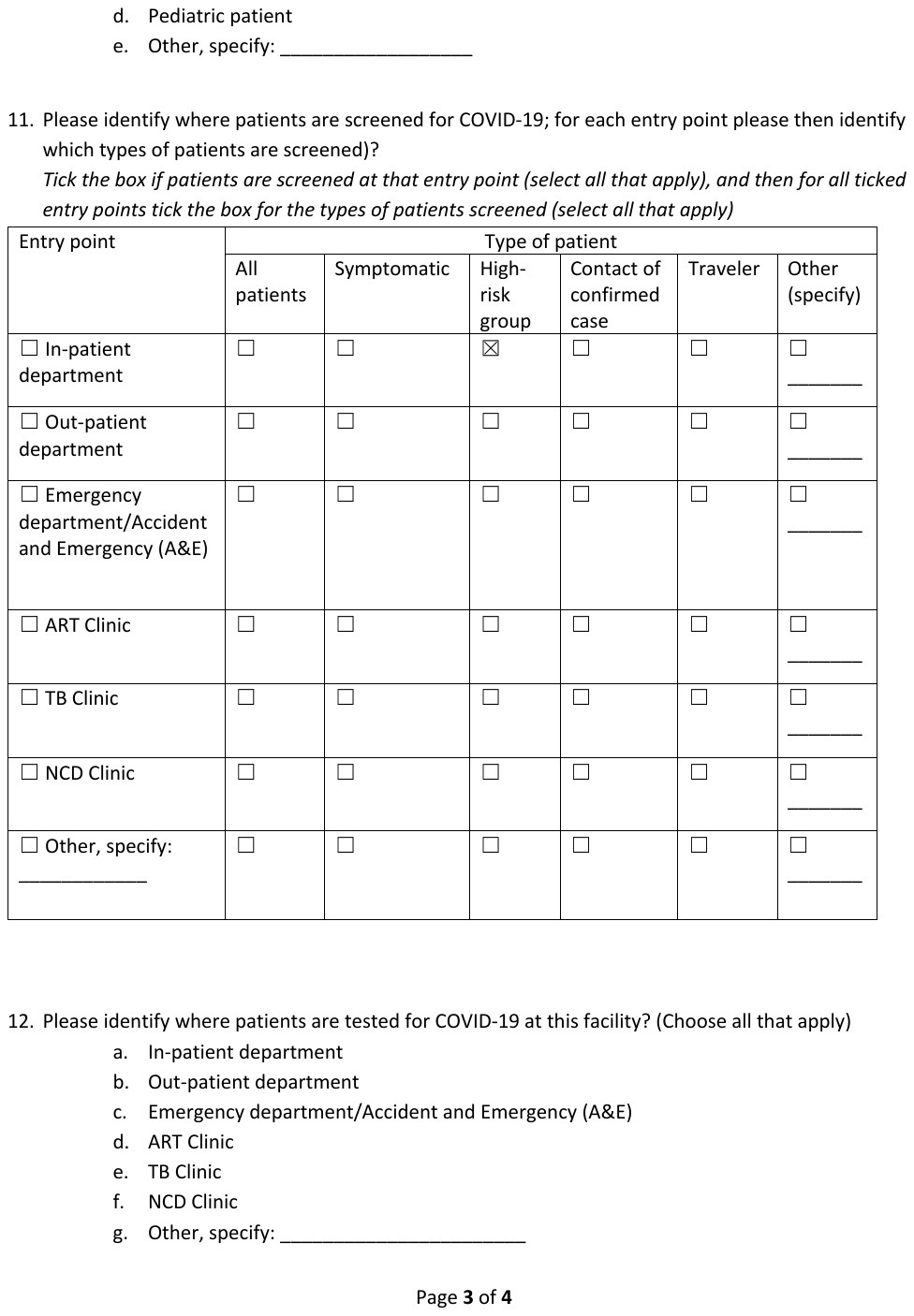

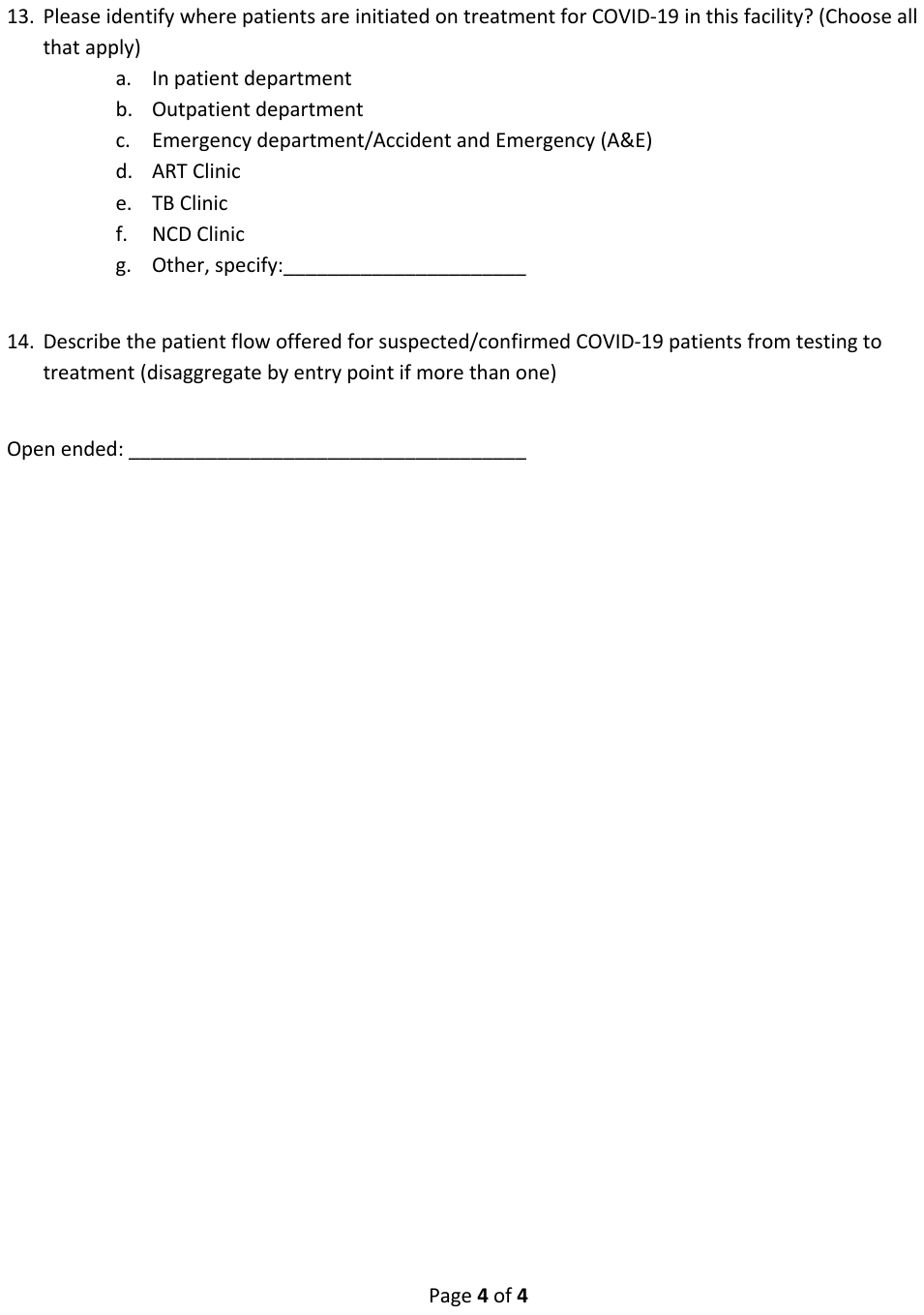

**Figure 3:Pre-Implementation Survey**

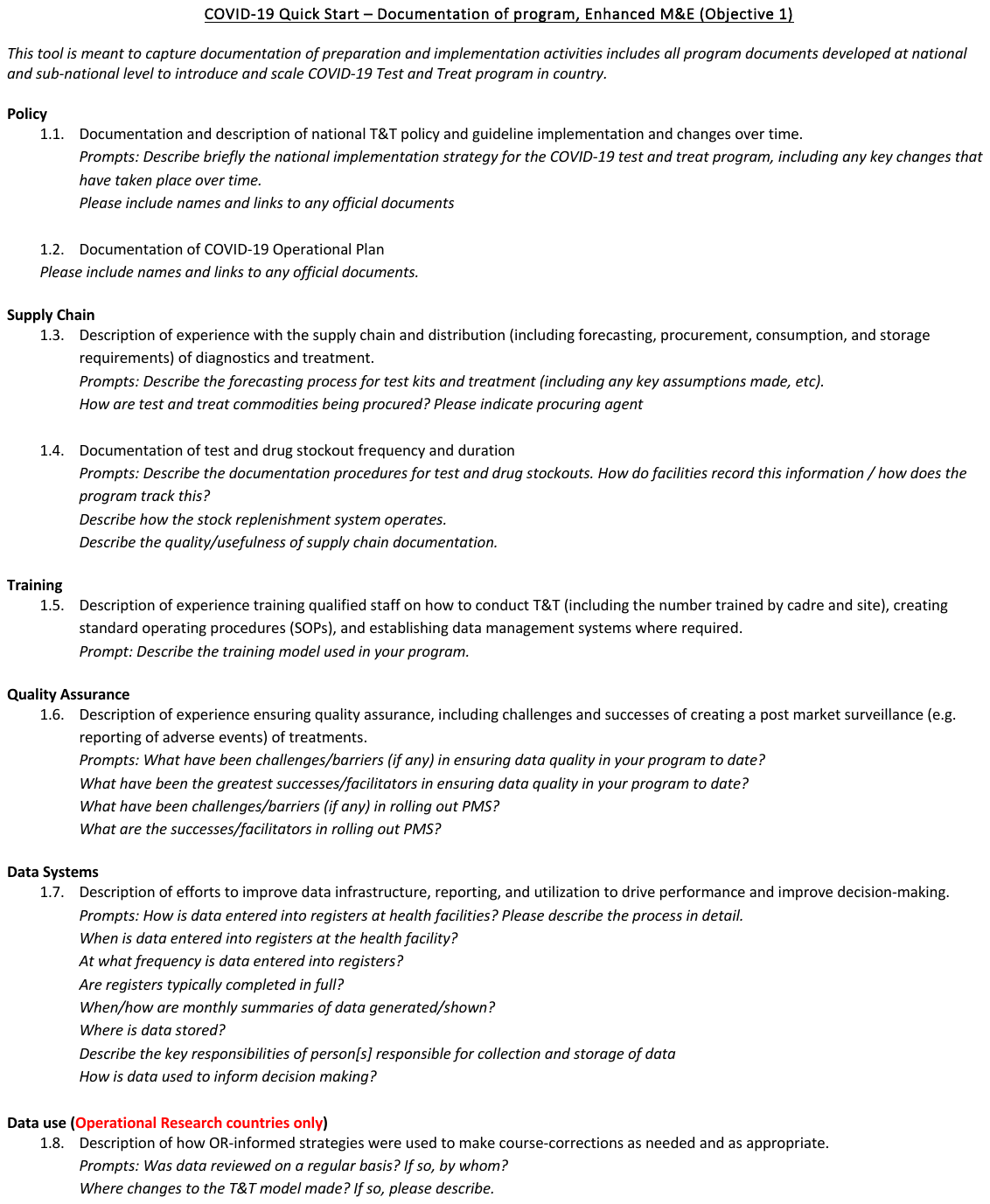

**Figure 4: MoH/Public Sector Personnel Interview Guide**

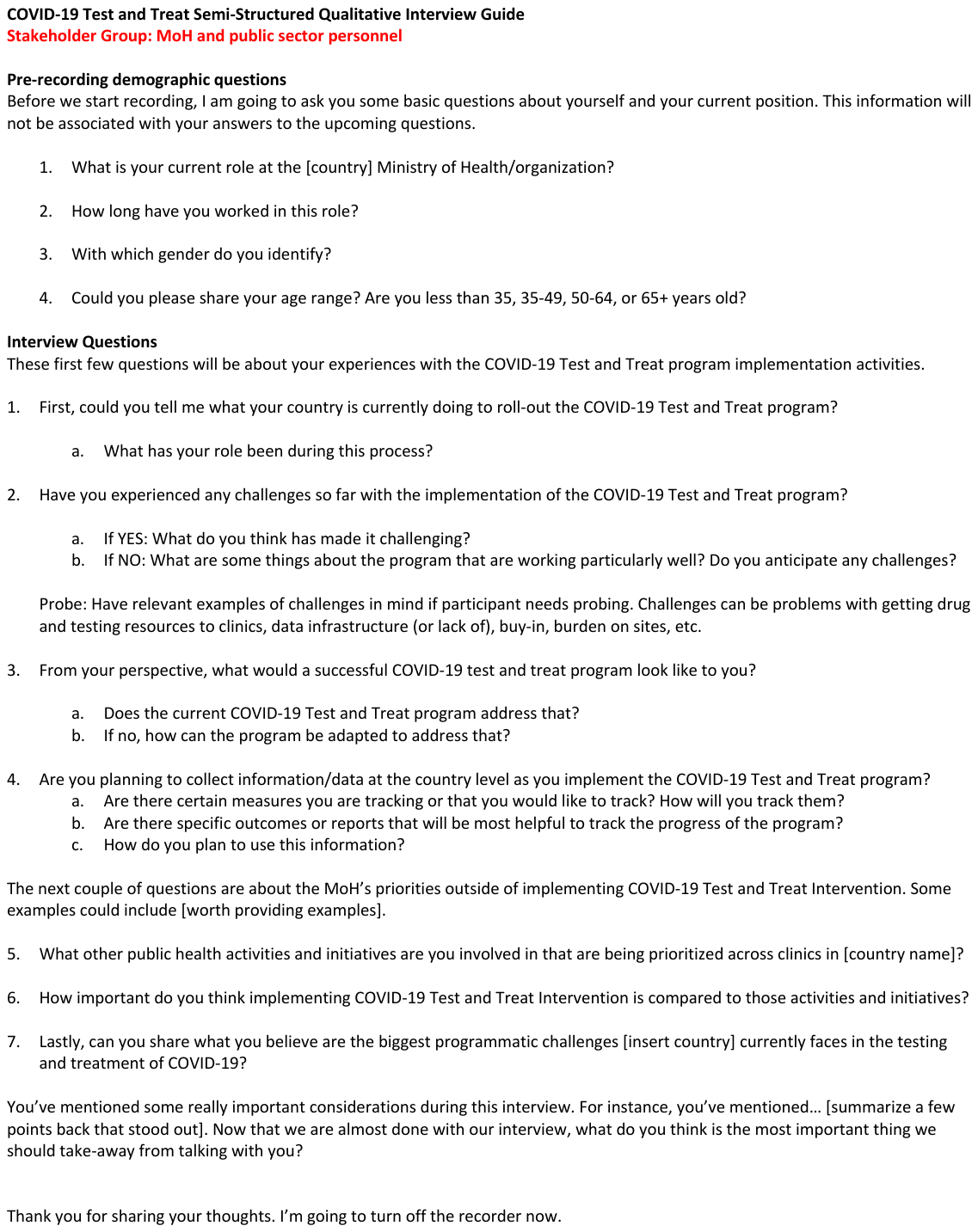

**Figure 5: Healthcare Worker Interview Guide**

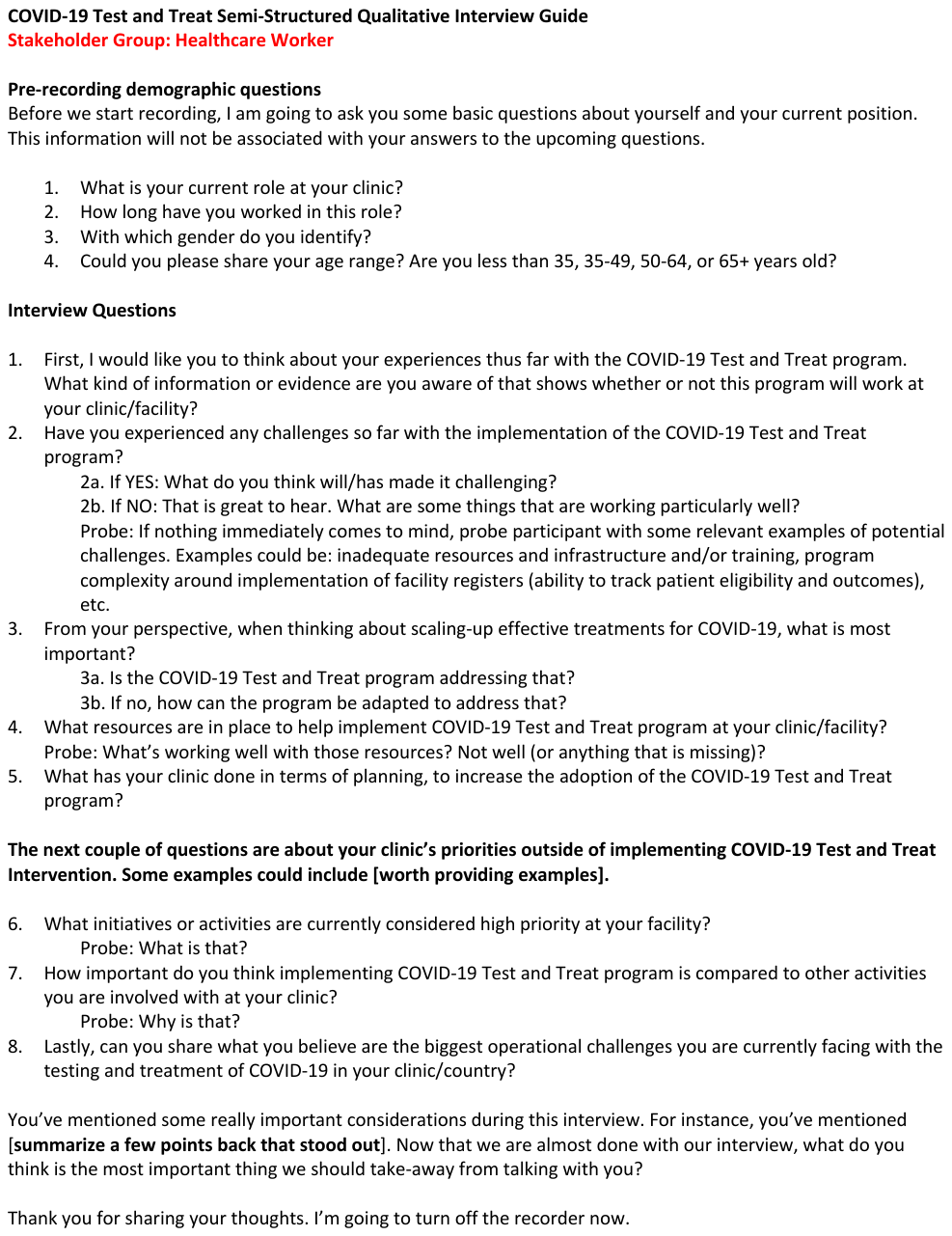

**Figure 6: Patient Interview Guide**

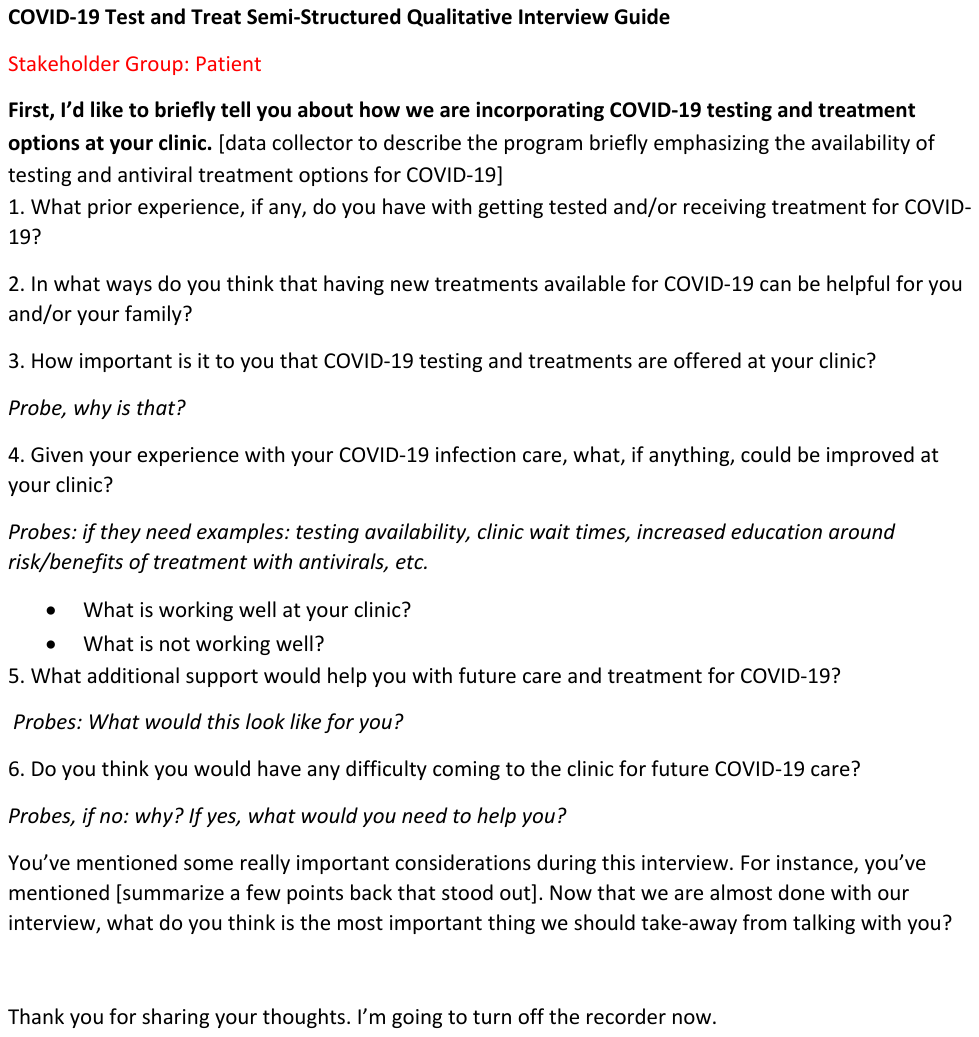
